# Interpreting chest X-rays via CNNs that exploit disease dependencies and uncertainty labels

**DOI:** 10.1101/19013342

**Authors:** Hieu H. Pham, Tung T. Le, Dat Q. Tran, Dat T. Ngo, Ha Q. Nguyen

## Abstract

Chest radiography is one of the most common types of diagnostic radiology exams, which is critical for screening and diagnosis of many different thoracic diseases. Specialized algorithms have been developed to detect several specific pathologies such as lung nodule or lung cancer. However, accurately detecting the presence of multiple diseases from chest X-rays (CXRs) is still a challenging task. This paper presents a supervised multi-label classification framework based on deep convolutional neural networks (CNNs) for predicting the risk of 14 common thoracic diseases. We tackle this problem by training state-of-the-art CNNs that exploit dependencies among abnormality labels. We also propose to use the label smoothing technique for a better handling of uncertain samples, which occupy a significant portion of almost every CXR dataset. Our model is trained on over 200,000 CXRs of the recently released CheXpert dataset and achieves a mean area under the curve (AUC) of 0.940 in predicting 5 selected pathologies from the validation set. This is the highest AUC score yet reported to date. The proposed method is also evaluated on the independent test set of the CheXpert competition, which is composed of 500 CXR studies annotated by a panel of 5 experienced radiologists. The performance is on average better than 2.6 out of 3 other individual radiologists with a mean AUC of 0.930, which ranks first on the CheXpert leaderboard at the time of writing this paper.

## 1. Introduction

Chest X-ray (CXR) is one of the most common radiological exams in diagnosing many different diseases related to lung and heart, with millions of scans performed globally every year [1]. Many diseases among them can be deadly if not diagnosed quickly and accurately enough. A computer-aided diagnosis (CAD) system that is able to interpret CXRs at a performance level comparable to practicing radiologists could provide substantial benefits for many realistic clinical contexts. In this work, we investigate the problem of multi-label classification for CXRs using deep convolutional neural networks (CNNs).

There has been a recent effort to harness advances in machine learning, especially deep learning, to build a new generation of CAD systems for classification and localization of common thoracic diseases from CXR images [2]. Several motivations are behind this transformation: First, interpreting CXRs to accurately diagnose pathologies is difficult. Even the best radiologists are prone to misdiagnoses due to challenges in distinguishing different kinds of pathologies, many of which often have similar visual features [3]. Therefore, a high-precision method for common thorax diseases classification and localization can be used as a second reader to support the decision making process of radiologists and to help reduce the diagnostic error. It also addresses the lack of diagnostic expertise in areas where the radiologists are limited or not available [4, 5]. Second, such a system can be used as a screening tool that helps reduce waiting time of patients in hospitals and allows care providers to respond to emergency situations sooner or to speed up a diagnostic imaging workflow [6]. Third, deep neural networks, in particular deep CNNs, have shown their remarkable performance for various applications in medical imaging analysis [7], including the CXR interpretation task.

Many deep learning-based approaches have been proposed for classifying lung diseases and proven that they could help radiologists overcome the limitations of human perception and bias, as well as reduce errors in diagnosis. Almost all of these approaches, however, aim to detect some specific diseases such as pneumonia [8], tuberculosis [9, 10], or lung cancer [11]. Meanwhile, building a unified deep learning framework for accurately detecting the presence of multiple common thoracic diseases from CXRs remains a difficult task that requires much research effort. In particular, we recognize that standard multi-label classifiers often ignore domain knowledge. For example, in the case of CXR data, how to leverage clinical taxonomies of disease patterns and how to handle uncertainty labels are still open questions, which have not received much research attention. This observation motivates us to build and optimize a predictive model based on deep CNNs for the CXR interpretation in which dependencies among labels and uncertainty information are taken into account during both the training and inference stages. Specifically, we develop a deep learning-based approach that puts together the ideas of *conditional training* [12] and *label smoothing* [13] into a novel training procedure for classifying 14 common lung diseases and observations. We trained our system on more than 200,000 CXRs of the CheXpert dataset [14]—one of the largest CXR dataset currently available, and evaluated it on a validation set containing 200 studies, which were manually annotated by 3 board-certified radiologists. The proposed method is also tested against the majority vote of 5 radiologists on the hidden test set of the CheXpert competition that contains 500 studies.

This study makes several contributions. First, we propose a novel training strategy for multi-label CXR classification that incorporates (1) a conditional training process based on a predefined disease hierarchy and (2) a smoothing policy for uncertainty labels. The benefits of these two key factors are empirically demonstrated through our ablation studies. Second, we train a series of state-of-the-art CNNs on frontal-view CXRs of the CheXpert dataset for classifying 14 common thoracic diseases. Our best model, which is an ensemble of various CNN architectures, achieves the highest area under ROC curve (AUC) score on both the validation set and test set of CheXpert at the time being. Specifically, on the validation set, it yields an averaged AUC of 0.940 in predicting 5 selected lung diseases: *Atelectasis* (0.909), *Cardiomegaly* (0.910), *Edema* (0.958), *Consolidation* (0.957) and *Pleural Effusion* (0.964). This model improves the baseline method reported in [14] by a large margin of 5%. On the independent test set, we obtain a mean AUC of 0.930. More importantly, the proposed deep learning model is on average more accurate than 2.6 out of 3 individual radiologists in predicting the 5 selected thoracic diseases when presented with the same data^1^.

The rest of the paper is organized as follows. Related works on CNNs in medical imaging and the problem of multi-label classification in CXR images are reviewed in Section 2. In Section 3, we present the details of the proposed method with a focus on how to deal with dependencies among diseases and uncertainty labels. Section 4 provides comprehensive experiments on the CheXpert dataset. Section 5 discusses the experimental results, some key findings and limitations of this research. Finally, Section 6 concludes the paper.

## 2. Related works

### 2.1. Deep learning in medical imaging

Recently, thanks to the increased availability of large scale, high-quality labeled datasets [15, 14, 16] and high-performing deep network architectures [17, 18, 19, 20], deep learning-based approaches have been able to reach, even outperform expert-level performance for many medical image interpretation tasks [21, 22, 23, 24]. Most successful applications of deep neural networks in medical imaging rely on CNNs, which were introduced in 1998 by LeCun et *al*. [25] and revolutionized in 2012 by Krizhevsky et *al*. [26]. State-of-the-art CNN models are rapidly becoming the standard for a wide range of applications in medical imaging such as detection, classification, and segmentation. They were applied successfully for lung cancer detection [27], pulmonary tuberculosis detection [9], skin cancer classification [28] and many others [7, 2].

### 2.2. Multi-label classification of CXRs

Multi-label classification is a common setting in CXR interpretation in which each input sample can be associated with one or several labels [29, 30]. Due to its important role in medical imaging, a variety of approaches have been proposed in the literature. For instance, Rajpurkar et *al*. [21] introduced CheXNet—a DenseNet-121 model that was trained on the ChestX-ray14 dataset [15], which achieved state-of-the-art performance on over 14 disease classes and exceeded radiologist performance on pneumonia using the F1 metric. Rajpurkar et *al*. [22] subsequently developed CheXNeXt, an improved version of the CheXNet, whose performance is on par with radiologists on a total of 10 pathologies of ChestX-ray14. Another notable work based on ChestX-ray14 was by Kumar et *al*. [31] who presented a cascaded deep neural network to improve the performance of the multi-label classification task. Closely related to our paper is the work of Chen et *al*. [12], in which they proposed to use the *conditional training* strategy to exploit the hierarchy of lung abnormalities in the PLCO dataset [32]. In this method, a DenseNet-121 was first trained on a restricted subset of the data such that all parent nodes in the label hierarchy are positive and then finetuned on the whole data.

Recently, the availability of very large-scale CXR datasets such as CheXpert [14] and MIMIC-CXR [16] provides researchers with an ideal volume of data (224,316 scans of CheXpert and more than 350,000 of MIMIC-CXR) for developing better and more robust supervised learning algorithms. Both of these datasets were automatically labeled by the same report-mining tool with 14 common findings. Irvin et *al*. [14] proposed to train a 121-layer DenseNet on CheXpert with various policies for handling the uncertainty labels. In particular, uncertainty labels were either ignored (`U-Ignore` policy) or mapped to *positive* (`U-Ones` policy) or *negative* (`U-Zeros` policy). On average, this baseline model outperformed 1.8 out of 3 individual radiologists with an AUC of 0.907 when predicting 5 selected pathologies on a test set of 500 studies. In another work, Rubin et *al*. [33] introduced DualNet—a novel dual convolutional networks that were jointly trained on both the frontal and lateral CXRs of MIMIC-CXR.

Experiments showed that the DualNet provides an improved performance in classifying findings in CXR images when compared to separate baseline (*i*.*e*. frontal and lateral) classifiers.

### 2.3. Key differences

Standard multi-label learning methods (*e*.*g. one-versus-all* or *one-versus-one* [29]) treat all labels independently. In many clinical contexts, however, there are significant dependencies between disease labels in which lung and cardiovascular pathologies are not an exception [34]. We believe that such dependencies should be exploited in order to improve the performance of the predictive models. In this paper, we adapt the conditional training approach of [32] to extensively train a series of CNN architectures for the hierarchy of the 14 CheXpert pathologies [14], which is totally different from that of PLCO [32]. Unlike previous studies, we also propose the use of the label smoothing regularization (LSR) [13] to leverage uncertainty labels, which, as experiments will later show, significantly improves the uncertainty policies originally proposed in [14].

## 3. Proposed Method

In this section, we present details of the proposed method. We first give a formulation of the multi-label classification for CXRs and the evaluation protocol used in this study (Section 3.1). We then describe a new training procedure that exploits the relationship among diseases for improving model performance (Section 3.2). This section also introduces the way we use the LSR to deal with uncertain samples in the training data (Section 3.3).

## 3.1. Problem formulation

Our focus in this paper is to develop and evaluate a deep learning-based approach that could learn from hundreds of thousands of CXR images and make accurate diagnoses of 14 common thoracic diseases from unseen samples. These 14 diseases include *Enlarged Cardiomediastinum, Cardiomegaly, Lung Opacity, Lung Lesion, Edema, Consolidation, Pneumonia, Atelectasis, Pneumothorax, Pleural Effusion, Pleural Other, Fracture, Support Devices*, and *No Finding*. In this multi-label learning scenario, we are given a training set 𝒟 = {(**x**^(*i*)^, **y**^(*i*)^); *i* = 1, …, *N*} that contains *N* CXRs; each input image **x**^(*i*)^ is associated with label **y**^(*i*)^ ∈ {0, 1}^14^, where 0 and 1 correspond to *negative* and *positive*, respectively. During the training stage, our goal is to train a CNN, parameterized by weights ***θ***, that maps **x**^(*i*)^ to a prediction **ŷ**^(*i*)^ such that the cross-entropy loss function is minimized over the training set 𝒟. Note that, instead of the softmax function, in the multi-label classification, the sigmoid activation function

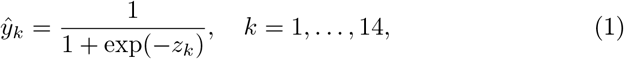

is applied to the logits *z*_*k*_ at the last layer of the CNN in order to output each of the 14 labels. The loss function is then given by

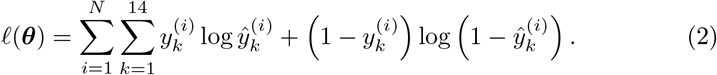

A validation set 𝒱 = {(**x**^(*j*)^, **y**^(*j*)^); *j* = 1, …, *M*} contains *M* CXRs, annotated by a panel of 5 radiologists, is used to evaluate the effectiveness of the proposed method. More specifically, model performance is measured by the AUC scores over 5 diseases: *Atelectasis, Cardiomegaly, Consolidation, Edema*, and *Pleural Effusion* from the validation set of the CheXpert dataset [14], which were selected based on clinical importance and prevalence. Figure 1 shows an illustration of the task we investigate in this paper.

**Figure 1:**
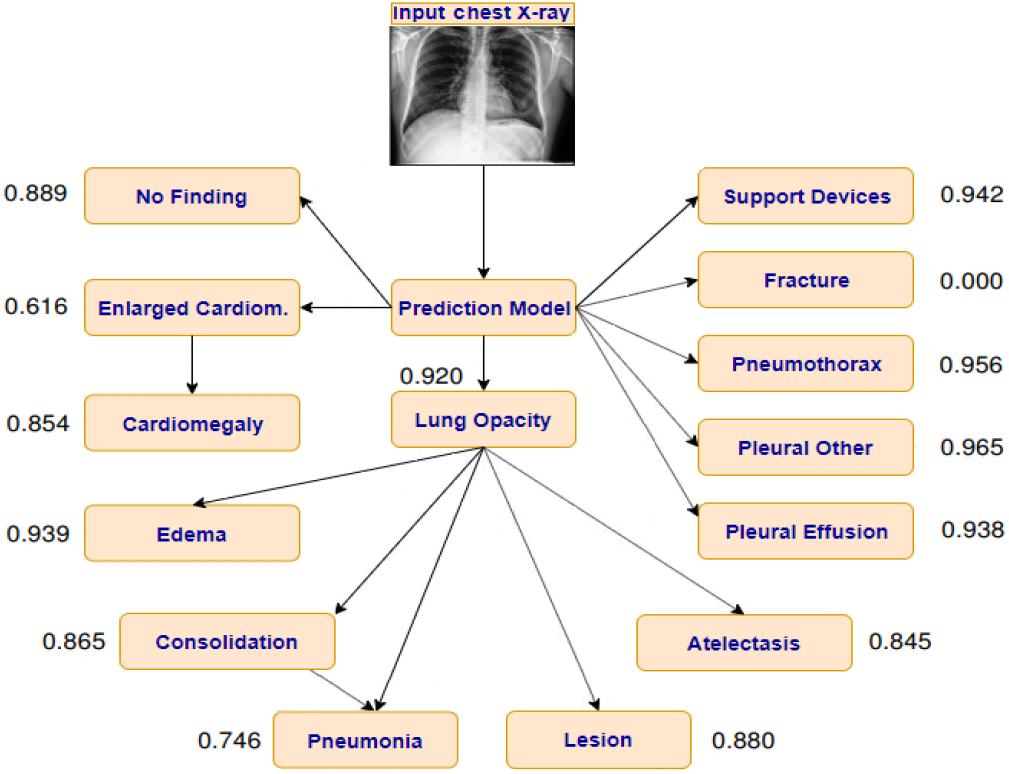
Illustration of our classification task, which aims to build a deep learning system for predicting probability of presence of 14 different pathologies or observations from the CXRs. The relationships among labels were proposed by Irvin et *al*. [14].

### 3.2. Conditional training to learn dependencies among labels

In medical imaging, labels are often organized into hierarchies in form of a tree or a directed acyclic graph (DAG). These hierarchies are constructed by domain experts, *e*.*g*. radiologists in the case of CXR data. Diagnoses or observations in CXRs are often conditioned upon their parent labels [34]. This important fact should be leveraged during the model training and prediction. Most existing CXR classification approaches, however, treat each label in an independent manner and do not take the label structure into account. This group of algorithms is known as *flat classification methods* [35]. A flat learning model reveals some limitations when applied to hierarchical data as it fails to model the dependency between diseases. For example, from Figure 1, the *Cardiomegaly* label is positive only if its parent, *Enlarged Cardiomediastinum*, is positive too. Additionally, some labels that are at the lower levels in the hierarchy, in particular at leaf nodes, have very few positive samples, which makes the the flat learning model more vulnerable to overfitting.

Another group of algorithms called *hierarchical multi-label classification methods* has been proposed for leveraging the hierarchical relationships among labels in making predictions, which has been successfully exploited for text processing [36], visual recognition [37, 38] and genomic analysis [39]. One common approach is to train classifiers on conditional data with all parent-level labels being positive and then to finetune them with the whole dataset [12], which contains both the positive and negative samples.

We adapt the idea of Chen *et al*. [12] to the lung disease hierarchy in Figure 1, which was initially introduced in [14]. Presuming the medical validity of the hierarchy, we break the training procedure into two steps. The first step, called *conditional training*, aims to learn the dependent relationships between parent and child labels and to concentrate on distinguishing lower-level labels, in particular the leaf labels. In this step, a CNN is pretrained on a partial training set containing all positive parent labels to classify the child labels; this procedure is illustrated in Figure 2. In the second step, *transfer learning* will be exploited.

**Figure 2:**
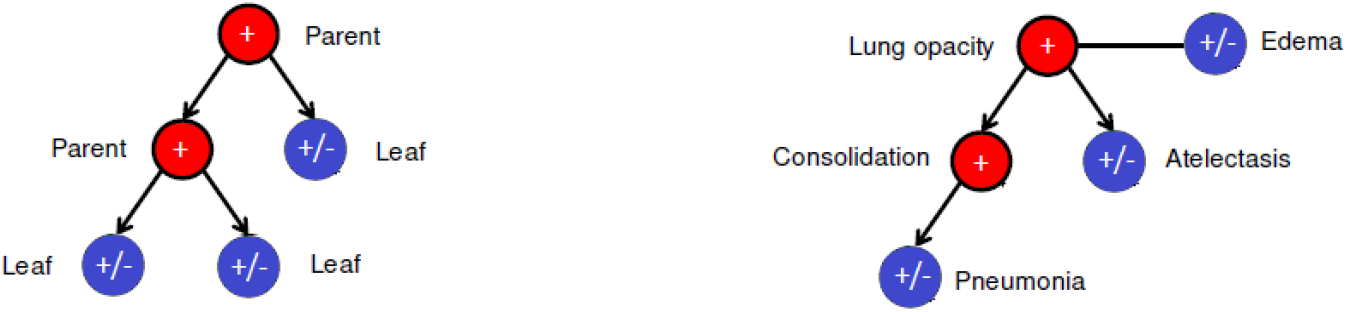
Illustration of the key idea behind the conditional training (**left**). In this stage, a CNN is trained on a training set where all parent labels (red nodes) are positive, to classify leaf labels (blue nodes), which could be either positive or negative. For example, we train a CNN to classify *Edema, Atelectasis*, and *Pneumonia* on training examples where both *Lung Opacity* and *Consolidation* are positive (**right**).

Specifically, we freeze all the layers of the pretrained network except the last fully connected layer and then retrain it on the full dataset. This training stage aims at improving the capacity of the network in predicting parent-level labels, which could also be either positive or negative.

According to the above training strategy, the output of the network for each label can be viewed as the conditional probability that this label is positive given its parent being positive. During the inference phase, however, all the labels should be unconditionally predicted. Thus, as a simple application of the Bayes rule, the unconditional probability of each label being positive should be computed by multiplying all conditional probabilities produced by the CNN along the path from the root node to the current label. For example, let 𝒞 and 𝒟 be disease labels at the leaf nodes of a tree 𝒯, which also parent labels 𝒜 andℬ, as drawn in Figure 3. Suppose the tuple of conditional predictions (*p*(𝒜), *p*(ℬ|𝒜), *p*(𝒞|ℬ), *p*(𝒟|𝒜)) are already provided by the network. Then, the unconditional predictions for the presence of 𝒞 and 𝒟 will be computed as

**Figure 3:**
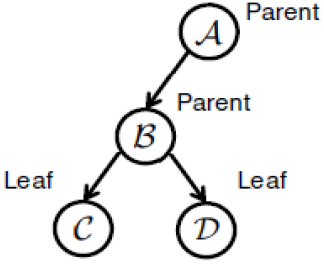
An example of a tree of 4 diseases: 𝒜, ℬ, 𝒞, and 𝒟.

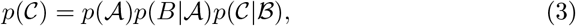

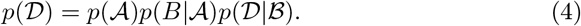

It is important to note that the *unconditional inference* mentioned above helps ensure that the probability of presence of a child disease is always smaller than the probability of its parent, which is consistent with clinical taxonomies in practice.

### 3.3. Leveraging uncertainty in CXRs with label smoothing regularization

Another challenging issue in the multi-label classification of CXRs is that we may not have full access to the true labels for all input images provided by the training dataset. A considerable effort has been devoted to creating large-scale CXR datasets with more reliable ground truth, such as CheXpert [14] and MIMIC-CXR [16]. The labeling of these datasets, however, heavily depends on expert systems (*i*.*e*. using keyword matching with hard-coded rules), which left many CXR images with uncertainty labels. Several policies have been proposed in [14] to deal with these uncertain samples. For example, they can be all *ignored* (`U-Ignore`), all mapped to *positive* (`U-Ones`), or all mapped to *negative* (`U-Zeros`). While `U-Ignore` could not make use of the whole dataset, both `U-Ones` and `U-Zeros` yielded a minimal improvement on CheXpert, as reported in [14]. This is because setting all uncertainty labels to either 1 or 0 will certainly produce a lot of wrong labels, which misguide the model training.

In this paper, we propose to apply a new advance in machine learning called *label smoothing regularization* (LSR) [40, 13, 41] for a better handling of uncertainty samples. Our main goal is to exploit the large amount of uncertain CXRs and, at the same time, to prevent the model from overconfident prediction of the training examples that might contain mislabeled data. Specifically, the `U-ones` policy is softened by mapping each uncertainty label (−1) to a random number close to 1. The proposed `U-ones+LSR` policy now maps the original label 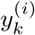 to

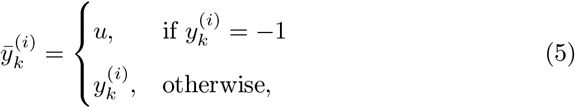

where *u* ∼ *U* (*a*_1_, *b*_1_) is a uniformly distributed random variable between *a*_1_ and *b*_1_—the hyper-parameters of this policy. Similarly, we propose the `U-zeros+LSR` policy that softens the `U-zeros` by setting each uncertainty label to a random number *u* ∼ *U* (*a*_0_, *b*_0_) that is closed to 0.

## 4. Experiments

### 4.1. CXR dataset and settings

CheXpert dataset [14] was used to develop and evaluate the proposed method. This is one of the largest and most challenging public CXR dataset currently available, which contains 224,316 scans of unique 65,240 patients, labeled for the presence of 14 common chest radiographic observations. Each observation can be assigned to either *positive* (1), *negative* (0), or *uncertain* (-1). The main task on CheXpert is to predict the probability of multiple observations from an input CXR. The predictive models take as input a single view CXR and output the probability of each of the 14 observations as shown in Figure 1. The whole dataset is divided into a training set of 223,414 studies, a validation set of 200 studies, and a test set of 500 studies. For the validation set, each study is annotated by 3 board-certified radiologists and the majority vote of these annotations serves as the ground-truth label. Meanwhile, each study in the test set is labeled by the consensus of 5 board-certified radiologists. The authors of CheXpert proposed an evaluation protocol over 5 observations: *Atelectasis, Cardiomegaly, Consolidation, Edema*, and *Pleural Effusion*, which were selected based on the clinical importance and prevalence from the validation set. The effectiveness of predictive models is measured by the AUC metric.

### 4.2. Data cleaning and normalization

The learning performance of deep neural networks on raw CXRs may be affected by the irrelevant noisy areas such as texts or the existence of irregular borders. Moreover, we observe a high ratio of CXRs that have poor alignment. We therefore propose a series of preprocessing steps to reduce the effect of irrelevant factors and focus on the lung area. Specifically, all CXRs were first rescaled to 256 × 256 pixels. A template matching algorithm [42] was then used to search and find the location of a template chest image (224 × 224 pixels) in the original images. Finally, they were normalized using mean and standard deviation of images from the ImageNet training set [26] in order to reduce source-dependent variation.

### 4.3. Network architecture and training methodology

We used DenseNet-121 [18] as a baseline network architecture for verifying our hypotheses on the conditional training procedure (Section 3.2) and the effect of LSR (Section 3.3). In the training stage, all images were fed into the network with a standard size of 224 × 224 pixels. The final fully-connected layer is a 14-dimensional dense layer, followed by sigmoid activations that were applied to each of the outputs to obtain the predicted probabilities of the presence of the 14 pathology classes. We used Adam optimizer [43] with default parameters *β*_1_ = 0.9, *β*_2_ = 0.999 and a batch size of 32 to find the optimal weights. The learning rate was initially set to 1*e*− 4 and then reduced by a factor of 10 after each epoch during the training phase. Our network was initialized with the pretrained model on ImageNet [26] and then trained for 5 epochs, which is equivalent to 50,000 iterations. During training, our goal is to minimize the binary cross-entropy loss function between the ground-truth labels and the predicted labels output by the network over the training samples. The proposed deep network was implemented in Python using Keras with TensorFlow as backend. All experiments were conducted on a Windows 10 machine with a single NVIDIA Geforce RTX 2080 Ti with 11GB memory.

We conducted extensive ablation studies to verify the impact of the proposed conditional training procedure and LSR. Specifically, we first trained independently the baseline network with 3 label policies: `U-Ignore`, `U-Ones`, and `U-Zeros`. We then fixed the hyperparameter settings of these runs above and performed the conditional training procedure on top of them, resulting in 3 other networks: `U-Ignore+CT`, `U-Ones+CT`, and `U-Zeros+CT`, respectively. Next, the LSR technique was applied to the two label policies `U-Ones` and `U-Zeros`. For `U-Ones`, all uncertainty labels were mapped to random numbers uniformly distributed in the interval [0.55, 0.85]. For `U-Zeros`, we labeled uncertain samples with random numbers in [0, 0.3]. Both of these intervals were emperically chosen. Finally, both CT and LSR were combined with `U-Ones` and `U-Zeros` using the same set of hyperparameters, resulting in `U-Ones+CT+LSR` and `U-Zeros+CT+LSR`, respectively.

### 4.4. Model ensembling

In a multi-label classification setting, it is hard for a single CNN model to obtain high and consistent AUC scores for all disease labels. In fact, the AUC score for each label often varies with the choice of network architecture. In order to achieve a highly accurate classifier, an ensemble technique should be explored. The key idea of the ensembling is to rely on the diversity of a set of possibly weak classifiers that can be combined into a stronger classifier. To that end, we trained and evaluated a strong set of different state-of-the-art CNN models on the CheXpert. The following architectures were investigated: DenseNet-121, DenseNet-169, DenseNet-201 [18], Inception-ResNet-v2 [19], Xception [44], and NASNetLarge [20]. The ensemble model was simply obtained by averaging the outputs of all trained networks. In the inference stage, the test-time augmentation (TTA) [45] was also applied. Specifically, for each test CXR, we applied a random transformation (amongst horizontal flipping, rotating ±7 degrees, scaling ±2%, and shearing ±5 pixels) 10 times and then averaged the outputs of the model on the 10 transformed samples to get the final prediction.

### 4.5. Quantitative results

Table 1 provides the AUC scores for all experimental settings we have conducted on the CheXpert validation set. We found that the best performing DenseNet-121 model was trained with the `U-Ones+CT+LSR` policy, which obtained an AUC of 0.894 on the validation set. This is a 4% improvement compared to the baseline trained with the `U-Ones` policy (mean AUC = 0.860). Additionally, experimental results show that both the proposed conditional training and LSR help boost the model performance. Our final model, which is an ensemble of six single models, achieved an average AUC of 0.940. As shown in Table 2, this score outperforms all previous state-of-the-art results. Figure 4 plots the ROC curves of the ensemble model for 5 pathologies on the validation set. Figure 5 illustrates some example predictions by the model during the inference stage.

**Table 1:** Experimental results on the CheXpert dataset measured by AUC metric over 200 CXR studies of the validation set. `CT` and `LSR` stand for the *conditional training* and *label smoothing regularization*, respectively. For each label policy, the highest AUC scores are boldfaced.

| Method | Atelectasis | Cardiomegaly | Consolidation | Edema | P. Effusion | Mean |
| --- | --- | --- | --- | --- | --- | --- |
| Ignore | 0.768 | 0.795 | 0.915 | **0.914** | 0.925 | 0.863 |
| Ignore+CT | **0.780** | **0.815** | **0.922** | **0.914** | **0.928** | **0.872** |
| U-Zeros | 0.745 | 0.813 | 0.882 | 0.921 | **0.930** | 0.858 |
| U-Zeros+CT | 0.782 | **0.835** | 0.922 | 0.923 | 0.921 | 0.877 |
| U-Zeros+LSR | 0.781 | 0.815 | 0.920 | 0.923 | 0.918 | 0.871 |
| U-Zeros+CT+LSR | **0.806** | 0.833 | **0.929** | **0.933** | 0.921 | **0.884** |
| U-Ones | 0.800 | 0.780 | 0.882 | 0.918 | 0.920 | 0.860 |
| U-Ones+CT | 0.813 | 0.816 | 0.895 | 0.923 | 0.912 | 0.872 |
| U-Ones+LSR | 0.818 | 0.834 | 0.874 | 0.925 | 0.921 | 0.874 |
| U-Ones+CT+LSR | **0.825** | **0.855** | **0.937** | **0.930** | **0.923** | **0.894** |

**Table 2:** Performance comparison using AUC metric with the state-of-the-art approaches on the CheXpert dataset. The highest AUC scores are boldfaced.

| Method | Atelectasis | Cardiomegaly | Consolidation | Edema | P. Effusion | Mean |
| --- | --- | --- | --- | --- | --- | --- |
| Ignore-LP [46] | 0.720 | 0.870 | 0.770 | 0.870 | 0.900 | 0.826 |
| Ignore-BR [46] | 0.720 | 0.880 | 0.770 | 0.870 | 0.900 | 0.828 |
| Ignore-CC [46] | 0.700 | 0.870 | 0.740 | 0.860 | 0.900 | 0.814 |
| Ignore [14] | 0.818 | 0.828 | 0.938 | 0.934 | 0.928 | 0.889 |
| U-Zeros [14] | 0.811 | 0.840 | 0.932 | 0.929 | 0.931 | 0.888 |
| U-Ones [14] | 0.858 | 0.832 | 0.899 | 0.941 | 0.934 | 0.893 |
| U-MultiClass [14] | 0.821 | 0.854 | 0.937 | 0.928 | 0.936 | 0.895 |
| U-SelfTrained [14] | 0.833 | 0.831 | 0.939 | 0.935 | 0.932 | 0.894 |
| Ours | **0.909** | **0.910** | **0.957** | **0.958** | **0.964** | **0.940** |

**Figure 4:**
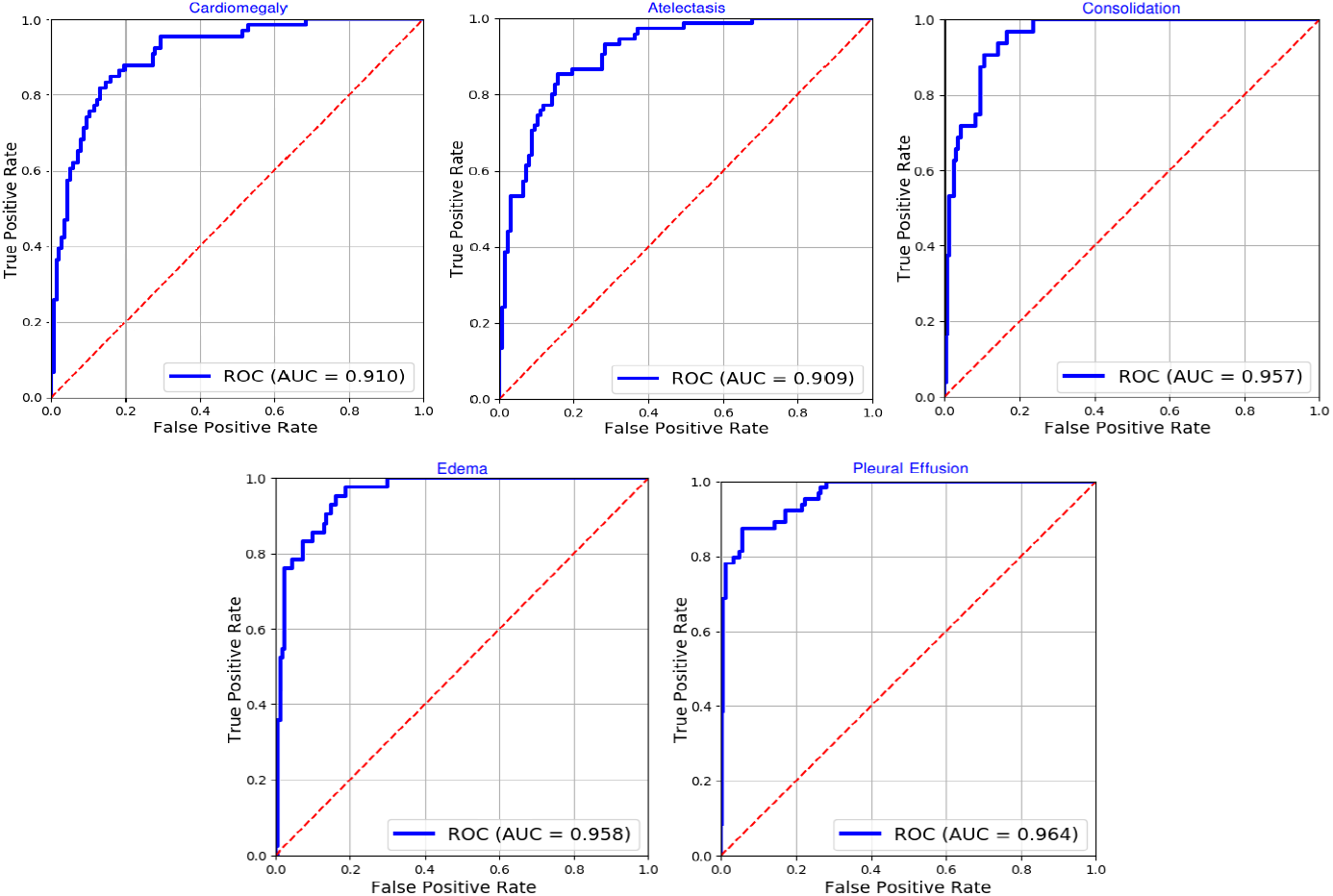
ROC curves of our ensemble model for the 5 pathologies on CheXpert validation set.

**Figure 5:**
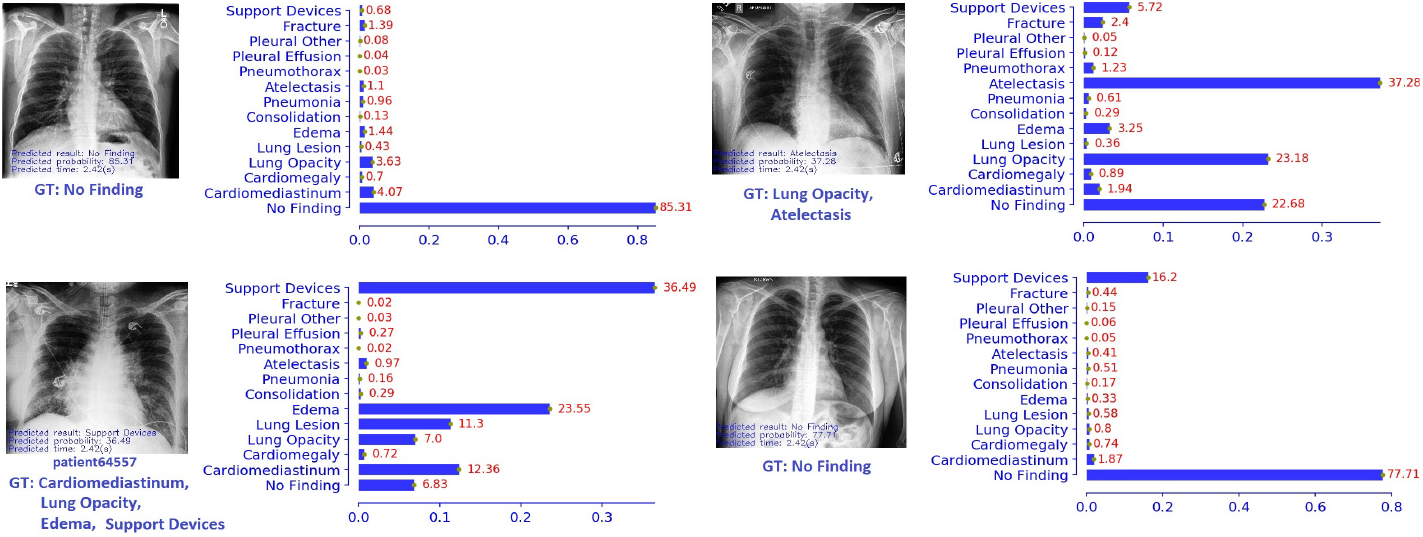
Visualization of findings by the proposed network during the inference stage.

### 4.6. Independent evaluation and comparison to radiologists

A crucial evaluation of any machine learning-based medical diagnosis system (ML-MDS) is to evaluate how well the system performs on an independent test set in comparison to human expert-level performance. To this end, we evaluated the proposed method on the hidden test set of CheXpert, which contains 500 CXRs labeled by 8 board-certified radiologists. The annotations of 3 of them were used for benchmarking radiologist performance and the majority vote of the other 5 served as ground truth. For each of the 3 individual radiologists, the AUC scores for the 5 selected diseases (*Atelectasis, Cardiomegaly, Consolidation, Edema*, and *Pleural Effusion*) were computed against the ground truth to evaluate radiologists’ performance. We then evaluated our ensemble model on the test set and performed ROC analysis to compare the model performance to radiologists. For more details, the ROCs produced by the prediction model and the three radiologists’ operating points were both plotted. For each disease, whether the model is superior to radiologists’ performances was determined by counting the number of radiologists’ operating points lying below the ROC^2^. The result shows that our deep learning model, when being averaged over the 5 diseases, outperforms 2.6 out of 3 radiologists with an AUC of 0.930. This is the best performance on the CheXpert leaderboard to date. The attained AUC score validates the generalization capability of the trained deep learning model on an unseen dataset. Meanwhile, the total number of radiologists under ROC curves indicates that the proposed method is able to reach human expert-level performance—an important step towards the application of an ML-MDS in real-world scenarios.

## 5. Discussions

### 5.1. Key findings and meaning

By training a set of strong CNNs on a large scale dataset, we built a deep learning model that can accurately predict multiple thoracic diseases from CXRs. In particular, we empirically showed a major improvement, in terms of AUC score, by exploiting the dependencies among diseases and by applying the label smoothing technique to uncertain samples. We found that it is especially difficult to obtain a good AUC score for all diseases with a single CNN. It is also observed that the classification performance varies with network architectures, the rate of positive/negative samples, as well as the visual features of the lung disease being detected. In this case, an ensemble of multiple deep learning models plays a key in boosting the generalization of the final model and its performance. Our findings, along with recent publications [21, 23, 22, 31], continue to assert that deep learning algorithms can accurately identify the risk of many thoracic diseases and is able to assist patient screening, diagnosing, and physician training.

### 5.2. Limitations

Although a highly accurate performance has been achieved, we acknowledge that the proposed method reveals some limitations. First, the deep learning algorithm was trained and evaluated on a CXR data source collected from a single tertiary care academic institution. Therefore, it may not yield the same level of performance when applied to data from other sources such as from other institutions with different scanners. This phenomenon is called *geographic variation*. To overcome this, the learning algorithm should be trained on data that are diverse in terms of regions, races, imaging protocols, etc. Second, to make a diagnosis from a CXR, doctors often rely on a broad range of additional data such as patient age, gender, medical history, clinical symptoms, and possibly CXRs from different views. This additional information should also be incorporated into the model training. Third, a finer resolution such as 512 × 512 or 1024 × 1024 could be beneficial for the detection of diseases that have small and complex structures on CXRs. This investigation, however, requires much more computational power for training and inference. Third, CXR image quality is another problem. When taking a deeper look at the CheXpert, we found a considerable rate of samples in low quality (*e*.*g*. rotated image, low-resolution, samples with texts, noise, etc.) that definitely hurts the model performance. In this case, a template matching-based method as proposed in this work may be insufficient to effectively remove all the undesired samples. A more robust preprocessing technique, such as that proposed in [47], should be applied to reject almost all *out-of-distribution* samples.

## 6. Conclusion

We presented in this paper a comprehensive approach for building a high-precision computer-aided diagnosis system for common thoracic diseases classification from CXRs. We investigated almost every aspect of the task including data cleaning, network design, training, and ensembling. In particular, we introduced a new training procedure in which dependencies among diseases and uncertainty labels are effectively exploited and integrated in training advanced CNNs. Extensive experiments demonstrated that the proposed method outperforms the previous state-of-the-art by a large margin on the CheXpert dataset. More importantly, our deep learning algorithm exhibited a performance on par with specialists in an independent test. There are several possible mechanisms to improve the current method. The most promising direction is to increase the size and quality of the dataset. A larger and high-quality labeled dataset can help deep neural networks generalize better and reduce the need for transfer learning from ImageNet. For instance, extra training data from MIMIC-CXR [16], which uses the same labeling tool as CheXpert, should be considered. We are currently expanding this research by collecting a new large-scale CXR dataset with radiologist-labeled reference from several hospitals and medical centers in Vietnam. The new dataset is needed to validate the proposed method and to confirm its usefulness in different clinical settings. We believe the cooperation between a machine learning-based medical diagnosis system and radiologists will improve the outcomes of thoracic disease diagnosis and bring benefits to clinicians and their patients.

## Data Availability

The dataset used in this research is available at https://stanfordmlgroup.github.io/competitions/chexpert/

## 7. Acknowledgements

This research was supported by the Vingroup Big Data Institute (VinBDI). The authors gratefully acknowledge Jeremy Irvin from the Machine Learning Group, Stanford University for helping us evaluate the proposed method on the hidden test set of CheXpert.

## Footnotes

1 Our model (Hierarchical-Learning-V1) currently takes the first place in the CheXpert competition. More information can be found at https://stanfordmlgroup.github.io/competitions/chexpert/. Updated on November 18, 2019.

2 This test was conducted independently with the support of the Stanford Machine Learning Group as the test set is not released to the public.

